# Haplotype of *RNASE 3* polymorphisms is associated with severe malaria in an Indian population

**DOI:** 10.1101/2020.06.16.20133090

**Authors:** Benudhar Mukhi, Himanshu Gupta, Samuel C. Wassmer, Anupkumar R. Anvikar, Susanta Kumar Ghosh

**Author notes:** Correspondence **Dr. Himanshu Gupta**, Department of Infection biology, Faculty of Infectious and Tropical Diseases, London School of Hygiene and Tropical Medicine, London, UK, Keppel St, Bloomsbury, London WC1E 7HT, Tel (M). Manipal Academy of Higher Education, Manipal – 576104, Karnataka, India (Benudhar Mukhi, Susanta Kumar Ghosh).

## Abstract

Severe malaria (SM) caused by *Plasmodium falciparum* (*Pf*) infection has been associated with life-threatening anemia, metabolic acidosis, cerebral malaria and multiorgan dysfunction. It may lead to death if not treated promptly. *RNASE 3* has been linked to *Pf* growth inhibition and its polymorphisms found associated with SM and cerebral malaria in African populations. This study aimed to assess the association of *RNASE 3* polymorphisms with SM in an Indian population. *RNASE 3* gene and flanking regions were amplified followed by direct DNA sequencing in 151 Indian patients who visited Wenlock District Government Hospital, Mangalore, Karnataka, India. Allele, genotype and haplotype frequencies were compared between patients with SM (n=47) and uncomplicated malaria (UM; n=104). Homozygous mutant genotype was only found for rs2233860 polymorphism (<1% frequency). No significant genetic associations were found for *RNASE 3* polymorphism genotypes and alleles in Indian SM patients. C-G-G haplotype of rs2233859, rs2073342 and rs2233860 polymorphisms was correlated significantly with SM patients (OR=3.03; p<0.001). Overall, these results suggest that *RNASE 3* gene plays a role in susceptibility to severe malaria.

## Introduction

Malaria remains to be a major public health problem in low and middle-income countries, especially in the sub-Saharan region. The World Health Organization (WHO) estimated that 228 million cases of malaria and 405,000 related deaths occurred globally in 2018 (WHO, 2019). Nineteen sub-Saharan African countries and India were responsible for carrying approximately 85% of the worldwide burden (WHO, 2019). *Plasmodium falciparum* (*Pf*) malaria is a complex disease with a wide spectrum of clinical manifestations ranging from uncomplicated (UM) to severe malaria (SM). SM is defined by life-threatening anemia, metabolic acidosis, cerebral malaria (CM), and multiorgan system involvement (Wassmer et al., 2015). Sequestration of *Pf*-parasitized erythrocytes within the microvasculature of vital organs in the human host is considered a key pathogenic event leading to SM (Miller et al., 2002; Dondorp AM, 2008). *P. falciparum* erythrocyte membrane protein 1 (PfEMP1) is encoded by the multicopy *var* gene family and mediates the sequestration of parasitized erythrocytes to host receptors (Rowe et al., 2009; Turner et al., 2013).

Both parasite and host factors have been identified as significant contributors to SM (Miller et al., 2002). This includes parasite genes, and *var* groups A, B, as well as domain cassette (DC) 8, DC11 and DC13 have been shown to be associated with SM (Lavstsen et al., 2012; Magallón-Tejada et al., 2016). Similarly, among host genes, polymorphisms in ICAM-1, CD36, TNF-α, IFN-γ, interleukin-1β, complement receptor-1 (CR-1), ABCB1 and ADORA2A have linked to the development of SM (Sinha et al., 2008; Olaniyan et al., 2016; Nasr et al., 2014; Gyan et al., 2002; Ouma et al., 2008; Panda et al., 2012; Gupta et al., 2017; Gupta et al., 2015). More recently, ribonuclease 3 (*RNASE 3*), which encodes eosinophil cationic protein (ECP), was found to increase susceptibility to SM (Adu et al., 2011; Diop et al., 2018; Kurtzhals et al., 1998; Waters et al., 1987). Indeed, SM patients had higher ECP levels and hypereosinophilia compared to UM patients (Kurtzhals et al., 1998). In addition, a parallel study demonstrated that ECP can suppress the growth of *Pf* in *in vitro* (Waters et al., 1987). These findings led to several genetic studies of the *RNASE 3* gene in African populations (Adu et al., 2011; Diop et al., 2018), which all showed an association between *RNASE 3* polymorphisms and SM (Adu et al., 2011; Diop et al., 2018), further confirming its role in severity of the disease.

Here, we conducted a case-control study to assess the association of *RNASE 3* polymorphisms with SM in India, as the same polymorphism can have heterogeneous effect in two populations due to genetic differences (Lin et al., 2007). *RNASE 3* polymorphism alleles, genotypes and haplotypes frequencies were compared between falciparum malaria patients with SM and UM.

## Materials and methods

### Ethical Statement

All subjects were recruited from the Department of Medicine, Wenlock District Government Hospital, Mangalore, Karnataka, India. Prior written informed consent was obtained from each adult patient, or informed assent from a parent or legal guardian if the individual was ≤ 18 years. The research and ethics committee of the Kasturba Medical College (KMC) under Manipal Academy of Higher Education, Mangalore, Karnataka, India, approved the study (IEC KMC MLR 03-16/49). The Institutional review board of ICMR-National Institute of Malaria Research, New Delhi, India also reviewed and approved the study (ECR/NIMR/EC/2012/39). Patient data obtained in this study was kept confidential and unique laboratory code was used for laboratory and dataset analyses.

### Study design and population

For this case-control study, we enrolled patients with SM and UM caused by *P. falciparum* who were admitted or visited to the Department of Medicine, Wenlock District Government Hospital, Mangalore, Karnataka, India, from July 2015 to December 2018. UM patients were used as controls to understand the role of *RNASE 3* polymorphisms in severe malaria. SM was defined based on the modified WHO criteria (WHO, 2015). A total of 151 patients were recruited (age range: 1-75 years), including 19 children (≤ 18 years, 12.6%) and 132 adults (>18, 87.4%) participants; 19 patients were female (12.6%). All the participants were from the same ethnic group, as determined on the basis of shared history, food habit, language and habitat region. Patients with axillary temperature >37.5°C and confirmed mono-infection of *Pf* by expert microscopy and rapid diagnostic tests (RDTs) were included in the study. *P. falciparum* positive patients but also had other *Plasmodium species*, HIV, HBsAg, HCV, pneumonia, bacterial meningitis, sepsis and tuberculosis infections were excluded from the study.

### Parasite quantification and treatment

Giemsa-stained thick and thin blood smears were air-dried, and tested for the presence of *Pf* parasites under a light microscope fitted with a 100X oil immersion lens and a 10X eyepiece (Zeiss Primo Star, Germany), and parasitemia was quantified as previously described (Punnath et al., 2019). In addition, the National Vector Borne Disease Control Programme (NVBDCP) approved RDT kits were used as per the manufacturer’s instructions to confirm *Plasmodium* infections. These kits were FalciVax™ Rapid Test for Malaria Pv/Pf (Ref. No.: 50301002), Onsite Malaria Pf/Pv Ag Rapid Test (Ref. No.: R0112C) and SD Bioline Malaria Ag P.f/P.v (Ref. No.: 05FK80) targeting both *P. vivax*-specific pLDH and *P. falciparum*-specific HRP-2 antigens. All the cases were successfully treated with the artemisinin-based combination therapy (ACT) as prescribed by the National Vector Borne Control Programme.

### Laboratory procedures

Patients positive for mono-*Pf* infections were subjected to venipuncture; 4ml blood was collected in EDTA Vacutainers (BD Vacutainer®) for hematological tests, and a further 4ml blood was taken for biochemical liver and kidney function tests using Clot Activator Vacutainers (BD Vacutainer®). DxH 800 Hematology (Beckman Coulter) and Cobas® 6000 (Roche) analyzers were used for hematological and biochemical tests, respectively.

### Molecular procedures

EDTA blood collected for each patient was sent to Chromous Biotech, Pvt. LTD, Bangalore, India (http://www.chromous.com/index.php?q=chromous-biotech/about-us), for DNA isolation, *RNASE 3* gene amplification and DNA Sanger sequencing. In brief, 100 - 250μl of blood was used for DNA isolation using the Chromous Biotech DNA extraction kit (Cat. No.: RKN25/26). Finally, DNA was eluted in 35μl of elution buffer available in the kit. *RNASE* 3 gene was amplified using two primer sets (**Set-1**: Fw: 5’-TCCAGCAAGAGTGGTGGATGAGAT-3’ and Rv: 5’-CTGTTGTCACATGCAACTACATAG-3’; **Set-2**: Fw: 5’-TTGCCATCCAGCACATCAGTCTGA and Rv: 5’-CTGGTTCCACCTCTATTACGATTGC-3’) covering 2047bp region (chr14: 20890932 - 20892978) of the *RNASE* 3 gene. In brief, *RNASE* 3 gene fragments were amplified separately in 50μl reactions including 200ng of each forward and reverse primers of set-1 and set-2, 5μl of 10x PCR buffer, 2μl of dNTPs (10mM), 2μl of template DNA and 3 units of Taq® DNA polymerase, reaction volume was raised by PCR-grade water. The reaction volume was prepared with PCR-grade water. The PCR reaction was performed with the initial denaturation at 94°C for 5min, followed by 35 cycles of 94°C for 30 sec, 52°C for 30 sec, 72°C for 2min, followed by final extension at 72°C for 7min. DNA sequencing (Sanger et al., 1977) was performed using ABI Prism (Applied Biosystems, USA) 3500 genetic analyzer automated DNA sequencer using ABI Prism BigDye Terminator v3.1 cycle sequencing kit. The direct DNA sequencing was performed for the two amplicons described above, of 1177bp and 1180bp long regions of *RNASE 3* containing all the reported gene polymorphisms. The variations in the sequences were identified by sequence alignment using NCBI blast with reference sequence NC_000014.9.

### Statistical analysis

To compare continuous data and categorical data between groups, respectively, we performed Mann Whitney U test and Fisher’s exact test. Odds ratio (OR), 95% confidence interval (95% CI) and p value for the mutant allele of each *RNASE 3* polymorphisms were calculated using online version of MedCalc software (https://www.medcalc.org/calc/odds_ratio.php). The SHEsis (http://analysis.bio-x.cn/SHEsisMain.htm), an online software tool, was used for haplotype and linkage disequilibrium (LD) analyses. All haplotypes with a frequency below 0.03 were discarded (Shi and He, 2005; Li et al., 2009). We defined statistical significance as p<0.05.

## Results

### Patient

All study participants (n=151) were positive for *P. falciparum* mono-infection, and no other species including *P. vivax, P. malariae, P. ovale* or *P. knowlesi* were identified. Among participants, median (± IQR) parasitemia was 39446 ± 52106 parasites/µL and median (± IQR) age was 27 ± 21years. Thrombocytopenia (<150,000 platelets/µL) was found in 60.9% of patients (92/151), and 10.6% (16/151) had severe thrombocytopenia (<50,000 platelets/µL). 31.1% (47/151) and 68.9% (104/151) of all patients were diagnosed with SM or UM, respectively. Comparisons between the demographic, hematological and biochemical laboratory findings of the participants with SM and UM was performed and are shown in **Table 1**. Statistically significant differences were only found for age, parasitemia, red blood cell counts, urea, bilirubin, AST, ALT, albumin levels and ratios of albumin and globulin (**Table 1**). Among patients with SM, 10.6% (5/47) and 23.4% (11/47) patients had multiple organ dysfunction and splenomegaly, respectively. Information on symptoms of severity of the patients as per the WHO criteria (WHO, 2015) is presented in **Table 2**.

**Table 1.** Demographic, hematological and biochemical laboratory findings of patients at the time of admission, 2015-2018. For continuous variables, values are expressed in median and interquartile range in bracket.

|  | SM (n=47) | UM (n=104) | p-value |
| --- | --- | --- | --- |
| **Female, no. (%)** | 6 (12.8) | 13 (12.5) | 1.000 |
| **Age (year)** | 36 (26.0) | 26 (20.0) | 0.007 |
| **Hemoglobin levels (g/dL)** | 12.3 (3.4) | 12.2 (3.5) | 0.828 |
| **Total RBC counts (million/mm<sup>3</sup>)** | 4.48 (1.1) | 4.92 (1.1) | 0.049 |
| **Platelet counts (per μL)** | 121000 (107000) | 112000 (65750) | 0.419 |
| **Blood glucose (mg/dL)** | 74 (37) | 79 (35.8) | 0.633 |
| **Blood urea (mg/dL)** | 37.5 (17.6) | 28.8 (16.1) | 0.002 |
| **Serum creatinine (mg/dL)** | 0.90 (0.7) | 0.88 (0.4) | 0.266 |
| **Serum bilirubin (mg/dL)** | 2.7 (2.8) | 1.6 (1.2) | < 0.001 |
| **AST levels (IU/L)** | 68.0 (61.5) | 56.7 (60.9) | 0.021 |
| **ALT levels (IU/L)** | 63.5 (40.5) | 41.5 (51.2) | 0.008 |
| **Alkaline phosphatase (IU/L)** | 310.4 (195.9) | 257.5 (198.7) | 0.092 |
| **Total protein levels (g/dL)** | 7.0 (1.3) | 7.1 (0.8) | 0.881 |
| **Albumin levels (g/dL)** | 3.7 (0.5) | 3.9 (0.7) | 0.038 |
| **Globulin levels (g/dL)** | 3.2 (1.2) | 3.1 (0.9) | 0.186 |
| **A:G Ratio** | 1.1 (0.4) | 1.2 (0.5) | 0.002 |
| **Parasitemia (per μL)** | 76357 (104280) | 28735 (40816) | < 0.001 |

*Severe malaria (SM); Uncomplicated malaria (UM); Albumin:globulin (A:G) ratio*

**Table 2.** Severe malaria symptoms according to WHO criteria in patients enrolled in the study.

| Severe malaria symptoms | Patients, n(%) |
| --- | --- |
| Metabolic acidosis | 30 (63.8) |
| Jaundice | 9 (19.1) |
| Hypoglycemia | 1 (2.1) |
| Severe anemia | 1 (2.1) |
| Acute kidney injury | 1 (2.1) |
| Pulmonary edema | 18 (38.3) |
| Multiple convulsions | 3 (6.4) |
| Hyperparasitemia | 1 (2.1) |

### Genetic association analyses

We successfully amplified 151 samples for *RNASE 3* gene followed by direct DNA sequencing. Among all the reported gene polymorphisms, homozygous mutant genotype was only found for rs2233860 polymorphism, only 0.7% (1/151) patients possessed a mutant genotype (CC). However, rs2233859 and rs2073342 polymorphisms were either present in the form of homozygous wild-type or heterozygous genotype in the studied participants **(Table 3)**. Thus, rs2233859, rs2073342 and rs2233860 were further considered for odds ratio, haplotype and linkage disequilibrium analyses.

**Table 3.**
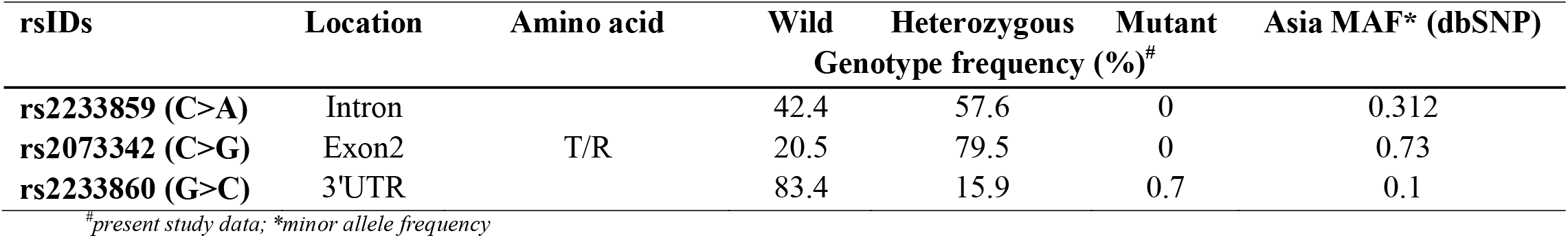
Genotype frequencies of studied polymorphisms in the Indian population.

Odds ratio was calculated for different genetic models, neither genotypes nor alleles of rs2233859, rs2073342 and rs2233860 polymorphisms were found associated with SM and UM patients (**Table 4)**.

**Table 4.** Odds ratio analysis of *RNASE 3* polymorphisms in severe and uncomplicated malaria patients.

| rsIDs | Genotypes/alleles | SM<br>(n=47) | UM<br>(n=104) | Models | OR (95%CI), p-value |
| --- | --- | --- | --- | --- | --- |
| rs2233859 | CC | 23 | 41 |  |  |
|  | CA | 24 | 63 |  |  |
|  | AA | 0 | 0 | Additive | NA |
|  | CA+AA | 24 | 63 | Dominant | 0.7 (0.3-1.3), p=0.274 |
|  | CC+CA | 47 | 104 | Recessive | NA |
|  | CC+AA | 23 | 41 | Co-dominant | 1.5 (0.7-2.9), p=0.274 |
|  | C | 70 | 145 |  |  |
|  | A | 24 | 63 | Allele | 0.8 (0.4-1.4), p=0.399 |
| rs2073342 | CC | 6 | 25 |  |  |
|  | CG | 41 | 79 |  |  |
|  | GG | 0 | 0 | Additive | NA |
|  | CG+GG | 41 | 79 | Dominant | 2.2 (0.8-5.7), p=0.118 |
|  | CC+CG | 47 | 104 | Recessive | NA |
|  | CC+GG | 6 | 25 | Co-dominant | 0.5 (0.2-1.2), p=0.118 |
|  | C | 53 | 129 |  |  |
|  | G | 41 | 79 | Allele | 1.3 (0.8-2.1), p=0.354 |
| rs2233860 | GG | 40 | 86 |  |  |
|  | GC | 7 | 17 |  |  |
|  | CC | 0 | 1 | Additive | NA |
|  | GC+CC | 7 | 18 | Dominant | 0.8 (0.3-2.2), p=0.712 |
|  | GG+GC | 47 | 103 | Recessive | NA |
|  | GG+CC | 40 | 87 | Co-dominant | 1.1 (0.4-2.9), p=0.821 |
|  | G | 87 | 189 |  |  |
|  | C | 7 | 19 | Allele | 0.8 (0.3-2.0), p=0.629 |

NA: not applicable due to the presence of 0 value; OR: odds ratio; 95%CI: 95% confidence interval; Severe malaria (SM); Uncomplicated malaria (UM)

The haplotype analysis was performed for three (rs2233859, rs2073342 and rs2233860) polymorphisms of *RNASE 3* gene by using SHEsis. Among the 8 possible haplotypes comprising the three polymorphic loci, C-G-G haplotype was associated with SM patients (OR=3.03; p<0.001). No, haplotypes were found associated with UM. The results of the haplotype analysis are shown in **Table 5**.

**Table 5.** Predicted haplotypes of *RNASE 3* polymorphisms (rs2233859, rs2073342 and rs2233860) association with severe and uncomplicated malaria.

| Haplotype | SM (frequency) | UM (frequency) | OR [95%CI] | Chi<sup>2</sup> | p value |
| --- | --- | --- | --- | --- | --- |
| A-C-C | 0.0 (0.0) | 0.18 (0.001) | - | - | - |
| A-C-G | 6.24 (0.066) | 5.92 (0.028) | 2.402 [0.759-7.603] | 2.35 | 0.125 |
| A-G-C | 0.0 (0.0) | 0.04 (0.0) | - | - | - |
| A-G-G | 17.76 (0.189) | 56.9 (0.274) | 0.610 [0.335-1.111] | 2.64 | 0.104 |
| C-C-C | 7.0 (0.074) | 16.84 (0.081) | 0.903 [0.361-2.261] | 0.05 | 0.827 |
| C-C-G | 39.76 (0.423) | 106.1 (0.510) | 0.690 [0.422-1.128] | 2.2 | 0.138 |
| C-G-C | 0.0 (0.0) | 1.98 (0.01) | - | - | - |
| C-G-G | 23.24 (0.247) | 20.12 (0.097) | 3.031 [1.572-5.845] | 11.66 | <0.001 |

OR: odds ratio; 95%CI: 95% confidence interval; Severe malaria (SM); Uncomplicated malaria (UM)

LD was estimated for the three polymorphisms using the calculated D′ values. The analysis showed strong linkage disequilibrium (D′ = 1 or > 0.75) between rs2233859, rs2073342 and rs2233860 polymorphisms in UM patients. However, no complete linkage between the three polymorphisms was observed in the SM group due to absence of LD between the polymorphisms rs2233859 and rs2073342 (D′ = 0.57). The results of the linkage analysis are shown in **Fig. 1**.

**Fig. 1.**
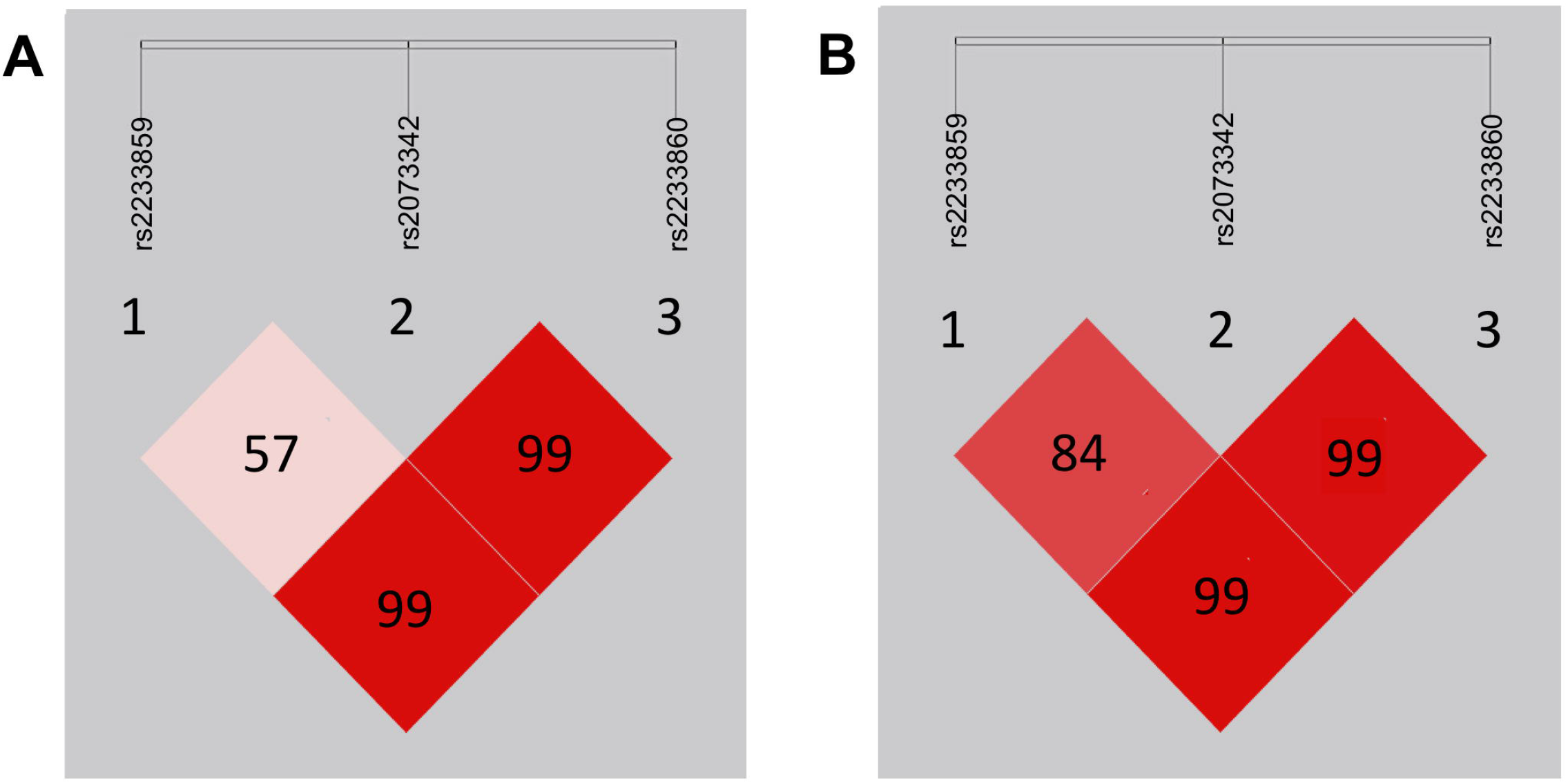
Linkage disequilibrium (LD) mapping of rs2233859, rs2073342 and rs2233860 polymorphisms of *RNASE 3* gene in patients with severe malaria (Panel A) and uncomplicated malaria (Panel B), the darkest shade indicating LD of 100 percent.

## Discussion

In this first study aiming to decipher the effect of *RNASE 3* polymorphisms on SM susceptibility in India, we used the direct DNA sequencing of clinical samples collected from *Pf*-positive patients residing in Mangalore, Karnataka, on the Western coast of India. We show evidence for the absence of homozygous mutant genotypes for *RNASE 3* polymorphisms, except for 3’ UTR rs2233860 polymorphism, which was only found in one patient. No significant genetic association was found for genotypes and alleles of *RNASE 3* polymorphisms with SM in the studied Indian population. This contrasts markedly with two reports on African populations, which showed an association between *RNASE 3* polymorphisms and SM susceptibility (Adu et al., 2011; Diop et al., 2018).

In Ghana, rs2073342 polymorphism was found associated with CM, a neurological type of SM, and rs2073342 and rs2233860 polymorphisms were associated with SM in a Senegalese population (Adu et al., 2011; Diop et al., 2018). No such associations were found in the present study. These differing results are may be due to several reasons. First, the same polymorphism can have heterogeneous effect in two geographically and genetically distinct populations, despite the endemicity of *P. falciparum* at both sites (Lin et al., 2007). Second, the present study did not include participants with CM and no past history of malaria, which could have provided higher granularity in our association study. Indeed, the SM patients enrolled in the Senegalese study consisted primarily of CM and severe anemia cases (Diop et al., 2018); it is therefore possible that the reported association was driven by a high number of CM patients, which would be in line with the findings from Ghana (Adu et al., 2011). It has to be noted that Mangalore city alone contributes about 72% of total malaria in Karnataka. In the last five years, the local district health department instead of the local municipal authority governs the malaria control operation. Following this, active and passive fever surveys through digital surveillance devices are in function throughout the city (Baliga et al., 2019). Thus prompt diagnosis and treatment is provided. This may be one of the reasons for not finding CM cases in the present study. However, further studies including large groups of different SM subtypes are needed to test this hypothesis in Indian populations.

We show that in SM patients from India, there is a lack of homozygous mutant genotypes of rs2073342 and rs2233860 polymorphisms. This absence may indicate that mutant allele may have a deleterious effect on the Indian population. This could be explained by the presence of heterozygous genotype in our cohort, may be to balance the deleterious effect of this mutant allele, especially for rs2073342 missense-polymorphism that has been associated with SM in both population of Ghana and Senegal (Adu et al., 2011; Diop et al., 2018). According to the dbSNP database, a 0.73 minor allele (G) frequency of rs2073342 polymorphism has been found in Asia **(Table 3)**, suggesting that it may increase in Indian population in the future.

Haplotype analyses can provide pivotal evidence on human evolution, and the identification of genetic variants causing specific human traits through linkage disequilibrium (Liu et al., 2008). C-G-G haplotype of rs2233859, rs2073342 and rs2233860 polymorphisms were found associated with SM (p<0.05) in this study. However, no complete linkage between the three polymorphisms was observed in the SM group. Therefore, in the absence of homozygous genotype for the mutant alleles of rs2233859 and rs2073342 polymorphisms, any conclusion on the association of C-G-G haplotype with SM would be speculative. Further genetic analyses involving a larger sample size in India are warranted to further explore the possible association we describe here.

SM predominantly affected adults in this study (median age 36 years, interquartile range 26 years), indicating a probable age shift in anti-malarial immunity (Fowkes et al., 2016). This may have resulted from the recent decrease in transmission due to India’s largest national malaria control program (Ghosh and Rahi, 2019). Higher bilirubin, AST and ALT levels were found in SM patients, confirming the association between hepatic injury and severe *Pf* infection described in previous studies (Anand et al., 1992; Chawla et al., 1989; Kochar et al., 2003a; 2003b). In addition, low albumin levels, albumin:globulin ratio, and high urea levels were noted in SM patients compared to UM, confirming the presence of liver and kidney injuries (**Table 2**). Parasitemia was also found significantly increased in SM patients. However, peripheral parasitemia may not be a true indicator of disease severity due to the sequestration of parasitized erythrocytes in the microvasculature of vital organs (Dondorp et al., 2005).

This study has a few limitations. In the era of next-generation sequencing and genome wide association studies, we performed a case-control study to assess the role of *RNASE 3* polymorphisms in SM susceptibility in Indian population using Sanger DNA sequencing. This option was chosen as case-control studies using Sanger sequencing assays are less time consuming, cost-effective, reliable, and can be useful for recognizing associating factors of disease outbreaks, as well as current cases, and allow the assessment of multiple risk factors at once (Tenny and Hoffman, 2020). Another limitation was the lack of CM patients included in the study, which may explain the discrepancies between our findings and the ones reported in Ghana and Senegal. Lastly, ECP levels were not assessed in plasma samples collected from study participants due to limited funding.

In conclusion, we have made an attempt to decipher the genetic association between *RNASE 3* polymorphisms and SM in an India population, and demonstrated that the homozygous mutant genotype of *RNASE 3* polymorphisms (rs2073342 and rs2233860), which were shown to be associated with CM and SM in the population of Ghana and Senegal, respectively, were absent in our cohort of SM patients. However, C-G-G haplotype of rs2233859, rs2073342 and rs2233860 polymorphisms were associated with a higher susceptibility to SM, further confirming a role of *RNASE 3* gene in malaria severity.

## Data Availability

Anonymized data is available on request

## Acknowledgment

We thank all study participants as well as physicians, technician, nurses from Wenlock District Government Hospital, Mangalore, Karnataka, India, and everyone who supported this study directly or indirectly. We thank the Director, National Institute of Malaria Research, New Delhi, for the opportunity to complete the research work. We also thank Dr. Rajeshwari Devi (District Surgeon and Medical Superintendent, Wenlock District Government Hospital, Mangalore) and Dr. K. Chakrapani (Associate Dean and Head of Medicine department) Kasturba Medical College (Manipal University), Mangalore, Karnataka, India, for their support on this study. We also acknowledge Indian Council of Medical Research, New Delhi, for funding. SCW is supported by a grant from the UK Medical Research Council, award number MR/S009450/1.

*Authors take this opportunity to applaud and thank health care workers all around the world for their hard work in the COVID-19 crisis*.

## Author contributions

BM participated in fieldwork, collected clinical and epidemiological data, laboratory analyses, and together with HG wrote the first draft of this manuscript. HG also participated in data curation, sequencing and statistical analyses as well as in results interpretation. SCW and AKRA participated in data interpretation and critically reviewing this article. SKG participated in the study design, supervision, generate the resources, manuscript review, project administration and coordinated all the stages of the project. All authors read and approved the final manuscript.

## Availability of data and material

Anonymized data is available on request

## Compliance with Ethical Standards

## Ethical approval

The research and ethics committee of the Kasturba Medical College (KMC) under Manipal Academy of Higher Education, Mangalore, Karnataka, India, approved the study (IEC KMC MLR 03-16/49). The Institutional review board of ICMR-National Institute of Malaria Research, New Delhi, India also reviewed and approved the study (ECR/NIMR/EC/2012/39).

## Informed consent

Prior written informed consent was obtained from each adult patient, or informed assent from a parent or legal guardian if the individual was ≤ 18 years.

